# Neural and visual processing of social gaze cueing in typical and ASD adults

**DOI:** 10.1101/2023.01.30.23284243

**Authors:** Termara Cerene Parker, Xian Zhang, Jack Adam Noah, Mark Tiede, Brian Scassellati, Megan Kelley, James Charles McPartland, Joy Hirsch

## Abstract

Atypical eye gaze in joint attention is a clinical characteristic of autism spectrum disorder (ASD). Despite this documented symptom, neural processing of joint attention tasks in real-life social interactions is not understood. To address this knowledge gap, functional-near infrared spectroscopy (fNIRS) and eye-tracking data were acquired simultaneously as ASD and typically developed (TD) individuals engaged in a gaze-directed joint attention task with a live human and robot partner. We test the hypothesis that face processing deficits in ASD are greater for interactive faces than for simulated (robot) faces. Consistent with prior findings, neural responses during human gaze cueing modulated by face visual dwell time resulted in increased activity of ventral frontal regions in ASD and dorsal parietal systems in TD participants. Hypoactivity of the right dorsal parietal area during live human gaze cueing was correlated with autism spectrum symptom severity: Brief Observations of Symptoms of Autism (BOSA) scores (r = −0.86). Contrarily, neural activity in response to robot gaze cueing modulated by visual acquisition factors activated dorsal parietal systems in ASD, and this neural activity was not related to autism symptom severity (r = 0.06). These results are consistent with the hypothesis that altered encoding of incoming facial information to the dorsal parietal cortex is specific to live human faces in ASD. These findings open new directions for understanding joint attention difficulties in ASD by providing a connection between superior parietal lobule activity and live interaction with human faces.

**Lay Summary:** Little is known about why it is so difficult for autistic individuals to make eye contact with other people. We find that in a live face-to-face viewing task with a robot, the brains of autistic participants were similar to typical participants but not when the partner was a live human. Findings suggest that difficulties in real-life social situations for autistic individuals may be specific to difficulties with live social interaction rather than general face gaze.

## Introduction

In face-to-face interactions of typically developed (TD) individuals, both adults and children focus their attention preferably and reflexively towards the direction of another person’s gaze. This two-way exchange of social information is defined as gaze-cueing of attention (Bayliss et al., 2007; Chen et al., 2021; Deaner & Platt, 2003). Gaze-cueing of attention facilitates joint attention, which involves the sharing of attention between two individuals toward the same object (Argyle & Cook, 1976; Baron-Cohen et al., 1985; Posner, 1980). In laboratory settings, a typical gaze cueing paradigm consists of a trial sequence in which a picture or a schematic face is presented centrally on a computer screen with eyes focusing in the direction of the observer (Itier & Batty, 2009). Subsequently, the gaze is shifted in the direction of a stimulus. The participant’s role is to detect, discriminate, or localize the target stimulus. Previous findings show that TD participants demonstrate significantly faster reaction times to detect a target location cued by the eye direction of the stimulus face than a non-cued target (Driver IV et al., 1999; Ristic & Kingstone, 2005). Gaze-directed attentional orientation is associated with neural responses in the dorsal parietal, right temporal parietal junction (rTPJ), and medial prefrontal cortices (Carlin & Calder, 2013; Colby & Goldberg, 1999; Dravida et al., 2020). Thus, gaze following involves both behavioral and neural processes that engage adaptive responses in TD individuals.

Autism spectrum disorder (ASD) is a neurodevelopmental disorder whose symptoms include atypical attentional responses to eye-gaze direction (Bagherzadeh-Azbari et al., 2022). Individuals with this condition are characterized by difficulties in social interaction and communication, as well as restrictive and repetitive behaviors (Maenner et al., 2020). Although gaze cueing studies indicate that the ability to follow gaze is intact in ASD (Nation & Penny, 2008), accumulating evidence demonstrates that autistic individuals distinguish less between social and non-social cues than their TD counterparts (Liu et al., 2022; Vlamings et al., 2005). These findings suggest that autistic individuals may possess distinct mechanisms for processing social gaze cues that differ from those employed by TD individuals. Furthermore, evidence from eye-tracking studies also shows that ASD individuals have difficulty with following the gaze of others (Freeth et al., 2010; Riby et al., 2013). These findings may reflect difficulties in understanding the “socialness” of the human eye gaze, subsequently leading to aberrant joint attention mechanisms in ASD (Freeth & Bugembe, 2019; Freeth et al., 2010). The process of actively sampling relevant social information using eye movements and fixations, visual sensing (Schroeder et al., 2010), is explored here as a contributing factor to joint attention difficulties in ASD (Sato & Uono, 2019).

In TD individuals, visual sensing allows for the holistic processing of faces, allowing rapid detection of changes in facial expression and eye gaze. Adaptations to facial expression and eye gaze are then coded at high levels of the facial information-processing hierarchy (Schroeder et al., 2010; Xu et al., 2008). Yet autistic individuals show both qualitative and quantitative differences in the processing of faces (Tang et al., 2015). Researchers have shown that autistic individuals may process cartoon faces easier than human faces (Cross et al., 2022; Rosset et al., 2010) as they may contain fewer dynamic facial expressions and eye movements. These findings provide a theoretical framework that predicts that autistic individuals may be better at engaging with simulated or cartoon faces than TD individuals. Additionally, the hierarchical specialization of different types of faces may differ between TD and ASD individuals.

Assistive robots, with cartoon-like faces, have been employed to understand and improve social functioning in ASD (Ismail et al., 2019; Pennisi et al., 2016; Scassellati et al., 2012). Autistic children show enhanced engagement with novel virtual and robotic partners, illustrating the potential value of these tools for intervention and therapy (Scassellati et al., 2012; Scassellati, Boccanfuso, et al., 2018). Studies have also shown increased spontaneous joint attention from children with ASD when engaging with virtual characters and robots (Cao et al., 2019; Porayska-Pomsta et al., 2012). Hou and colleagues suggest that the right dorsolateral prefrontal cortex is important in guiding young autistic children’s interest in robots in social contexts (Hou et al., 2022). Further knowledge of how ASD adults process robot-facilitated and human-facilitated joint attention will inform the future development of technological interventions as well as models of gaze-directed joint attention.

Classic gaze-cueing studies that use static robot or human faces on a computer screen contribute to the understanding of the cognitive and neural mechanisms of joint attention, but lack the aspect of reciprocity in live social interactions (Schilbach, 2015). Despite the social significance of assistive robots and known atypical responses to gaze direction, the underlying neural processes of live human- and robot-facilitated gaze-cueing in ASD have not been adequately investigated, due to limitations of conventional neuroimaging methods that fail to capture natural two-person interactions. Functional near-infrared spectroscopy (fNIRS) allows the assessment of neural responses under naturalistic conditions (Hirsch et al., 2018; Hirsch et al., 2017; Piva et al., 2017). Here, we investigate how adults with and without ASD respond to gaze cueing with a human and robot partner using simultaneous fNIRS and eye-tracking. We have previously shown that direct eye-to-eye contact and joint attention engage dorsal parietal systems during social interaction (Dravida et al., 2020; Hirsch et al., 2022; Hirsch et al., 2017; Kelley et al., 2021; Noah et al., 2020). Based on these prior findings, we hypothesize that encoding of oculomotor mechanisms such as face dwell time will be altered and associated with hypoactivity of the right superior parietal lobule and temporoparietal junction system for live human gaze cueing in ASD. If this effect is “human interactive specific,” then in the case of robot gaze cueing, we predict that the neural encoding of live face interaction will activate the superior parietal lobule in ASD participants, demonstrating that alterations to the typical dorsal parietal circuit are specific to live human faces and not faces in general. Furthermore, we examine how these neural responses relate to social symptomatology. If, as predicted, reductions in dorsal stream neural activity during human gaze cueing are correlated with autism symptom severity, then findings would suggest a possible biomarker for ASD.

## Methods and Materials

### Participants

Twenty ASD adults (Mean age 27±5.9 years; 18 right-handed, 2 left-handed (Oldfield, 1971) and 30 typically-developed (TD) adults (Mean age 23±4.4 years; 27 right-handed and 3 left) participated in this study (**Table 1**). ASD diagnoses were confirmed by gold standard, research-reliable clinician assessments, including the Autism Diagnostic Observation Schedule, 2nd Edition (ADOS-2 (Lord et al., 2012)), Brief Observation of Symptoms of Autism (BOSA (Lord et al., 2020)), and expert clinical judgment using DSM-5 criteria (American Psychiatric Association, 2013). Average ADOS-2 and BOSA Comparison Scores were 7 ± 0.26 and 7 ± 0.42, respectively. Assessment and diagnostic tests were performed in clinical facilities at the Yale Child Study Center and the Brain Function Laboratory. Participants were age-matched (**Table 1**) and recruited from ongoing research in the McPartland Lab, the Yale Developmental Disabilities Clinic, and the broader community through flyers and social media announcements. Inclusion criteria included age 18-45 years, IQ≥70, and English speaking. Exclusion criteria were the same as a previous investigation from the lab (Hirsch et al., 2022). All participants provided written and verbal informed consent under guidelines and regulations approved by the Yale University Human Investigation Committee (HIC # 1512016895) and were compensated for their participation. Assessment of the ASD participants’ capacity to give informed consent was provided by clinical research staff who monitored the process and confirmed verbal and non-verbal responses. ASD participants were accompanied by a member of the clinical team, who continuously evaluated their sustained consent to participate. Further information about participant demographics is outlined in Supplementary Methods.

**Table 1.** Participant Demographics

| TD Demographics |  |  |
| --- | --- | --- |
| **Gender** | n | % |
| Woman | 19 | 63.3% |
| Man | 10 | 33.3% |
| Non-Binary* | 1 | 3.3% |
|  | **30** | 100% |
| **Race** |  |  |
| Black | 2 |  |
| White | 13 |  |
| Asian | 13 |  |
| Latinx | 1 |  |
| Biracial | 1 |  |
| **Age (years)** |  |  |
| Mean | **23.8** |  |
| Median | 22 |  |
| Range | 18-35 |  |
| SD | ± 4.4 |  |
| ASD Demographics |  |  |

**Table 1.**
| **Gender** | n | % |
| --- | --- | --- |
| Woman | 3 | 15% |
| Man | 16 | 80% |
| Non-Binary* | 1 | 5% |
|  | **20** | 100% |
| **Race** |  |  |
| Black | 0 |  |
| White | 18 |  |
| Asian | 2 |  |
| Latinx | 0 |  |
| Biracial | 0 |  |
| **Age (years)** |  |  |
| Mean | **27.5** |  |
| Median | 25.6 |  |
| Range | 18-39 |  |
| SD | ± 6.0 |  |

\*self-identified gender is non-binary

A research investigator was present during data acquisition while monitoring signs of discomfort during the experiment. Each participant was paired with a TD initiator. Two females (22-23 years old throughout data collection) served as human initiators throughout the entire study. Determination of a sample size sufficient for a conventional power of 0.80 is based on contrasts (Real Face > Video Face) observed from a previous similar study (Noah et al., 2020). Using the “pwr” package of R statistical software (Champely et al., 2017), a significance level of p ≤ 0.05, uncorrected, is achieved with n of 16 subjects (for each diagnostic group). This estimate of sample size is consistent with similar calculations based on the signal strength in other relevant peak ROIs. Sample sizes of 20 participants (ASD) and 30 participants (TD) ensured adequate effect sizes. The gender composition of the ASD group is consistent with the estimated 4:1 male:female ratio of ASD diagnosis.

### Experimental Design and Statistical Analyses

The experiment consists of two conditions: gaze cueing with a human dyad partner who performs cue initiation and with a robot that has a simplified face that initiates the cue. Maki (HelloRobot, Atlanta, Georgia; (Payne, 2018); (Scassellati, Brawer, et al., 2018) is a robot capable of dynamic eye and head movements and was donated by Yale University’s Department of Computer Science. Participants engaged in a gaze cueing task in which an initiator (robot or human) used eye gaze to direct the participant’s eyes to one of two circular targets located either 13.4° to the left or right on an electronically controlled glass partition. Participants were directed to gather information from the face of the initiator to direct gaze to a specific location. The order of right and left directions was randomized.

Participants were seated at a table across from each other, approximately 140 cm. Between them, on the table, was the electronically controlled glass partition, or Smart Glass, that changed between transparent and opaque states (**Figure 1A**). The Smart Glass was pre-programmed to be transparent during the task blocks and opaque during rest periods. A scene camera was positioned on a camera mount attached to an articulated arm behind each participant and was aimed to record the participant’s view during the experiment. A human initiator received a visual cue (through a small screen on the other side of the glass not visible to the participant) during the rest periods. The robot initiator was electronically programmed to make movements directed to the target and to blink randomly. Once the Smart Glass changed from opaque to transparent, the initiator looked at the participant’s eyes for 2-seconds and then averted gaze to a target (left or right) on the glass partition for 2-seconds (**Figure 1B**). The participant’s task was to follow the initiator’s gaze. The paradigm consisted of 3 gaze cueing events followed by 15 seconds of rest (opaque Smart Glass) for a total of 3 minutes per run (**Figure 1C**). Eye-tracking was used to confirm participant compliance for each trial.

**Figure 1.**
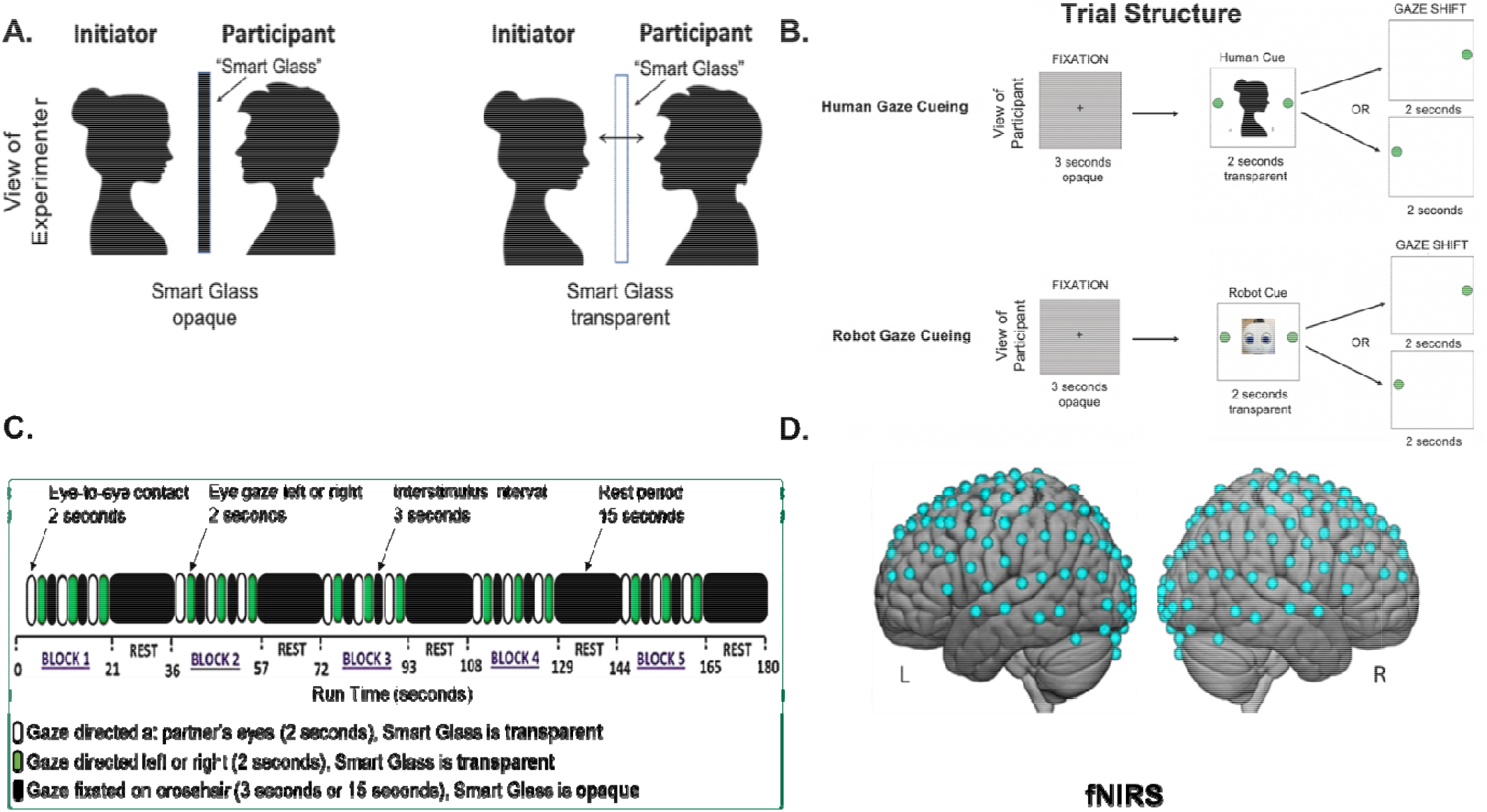
Gaze Cueing Tasks. **(A)** Schematic representation of the electronically controlled Smart Glass for both conditions. **(B)** Partners viewed each other at an eye-to-eye distance of 140 cm. Smart Glass is transparent for eye-to-eye contact (2s) and diverted gaze (2s). Smart Glass is opaque during interstimulus intervals (ISI, 3s) and rest periods (15 s). (**C**) Paradigm time course. Green and white bars represent 2-s trials. Task blocks consisted of three 4-s trials interspersed with 3-s of rest. 21-s task blocks alternated with 15-s rest blocks. (**D**) Example fNIRS 134-channel layout covering full cortical areas on one participant. Montreal Neurological Institute (MNI) coordinates were acquired for each channel (blue dots) by digitizing emitter and detector locations in relation to anterior, posterior, dorsal, and lateral fiduciary markers. Each marker was based on the standard 10-20 system.

### Eye-Tracking

Eye-tracking data for each participant was measured using a Tobii Pro x3-120 eye tracker (Tobii Pro, Stockholm, Sweden) at a sampling rate of 120 Hz, mounted on the experimental apparatus facing the participant. A three-point calibration method was utilized to calibrate the eye tracker on each participant before experimental recording. The initiator looked straight ahead while the participant was instructed to look at the eyes of the initiator, and then at three dot positions in front of the initiator’s face. The same calibration procedure for robot interactions was performed. Similar “live calibration” procedures have been used successfully in prior investigations of in-person social attention (Dravida et al., 2020; Falck-Ytter, 2015; Noah et al., 2020; Thorup et al., 2016). Participants interchanged their gaze between ≈0° and ±13.4° of deflection as instructed for the gaze cueing task. The eye contact portions of the task were 4 seconds in length, with 3 per trial, for 21 seconds of expected eye contact over the trial duration.

### Functional NIRS Signal Acquisition, Channel Localization, and Signal Processing

Similar methods have been previously described in other studies in our lab (Hirsch et al., 2022; Kelley et al., 2021; Noah et al., 2020) and detailed methods are described in Supplementary Methods. The specific layout with the coverage of the optode channels is shown in **Figure 1D**.

### Eye-tracking Analysis

Eye-tracking data were exported from the Tobii system to a custom data processing pipeline in MATLAB (Mathworks, Natick, MA). The MATLAB data processing pipeline calculated eye contact events, accuracy, and pupil diameter. One out of the 30 TD participants and two out of the 20 ASD participants did not provide usable eye-tracking data due calibration errors. Tobii Pro Lab software (Tobii Pro, Stockholm, Sweden) was used. For each run and each participant, a face box was manually defined for both human and robot gaze cueing conditions. For the visual sensing measures of gaze duration (Dwell Time) and Gaze Variability, the horizontal components of gaze trajectories from the gaze cue portions of each run were analyzed, focusing on the samples within the face box range. Dwell Time was determined by the number of valid retained samples per interval normalized by the sampling rate (seconds). Gaze Variability was computed as the standard deviation of the sample durations centered within the eye box.

### Participant Compliance

We asked our participants to follow the eyes of the initiator during the cued 4-seconds and compliance was measured by eye-tracking. For the TD group, eye following accuracy was 100% in both the robot and human gaze cueing conditions. For the ASD group, eye following accuracy was 100% in the robot condition and 99.4% for the human gaze cueing condition (**Supplementary Figure 1**).

## Results

### Visual Sensing Findings

The first two seconds of the event represent the time spent gathering information from the human or robot face before directing gaze to a target location. The recorded measures of dwell time in the “human face box” during the initial two second period did not differ between TD and ASD groups for the human initiator conditions [TD: 1.6943 ± 0.0288 seconds; ASD: 1.6067 ± 0.0688 seconds; t=1.3056, p=0.1983] (**Figure 2A)**. Similarly, gaze variability, the standard deviation of horizontal trajectory normalized by the duration of human face contact, also did not differ statistically between groups [TD: 0.0092 ± 0.0024; ASD: 0.0220 ± 0.0088; t=-1.6549; p=0.1049] (**Figure 2B**). However, average dwell time in the “robot face box” during the first two seconds of each robot gaze cueing trail was longer for the TD group [TD: 1.7479 ± 0.0205 seconds; ASD: 1.5745 ± 0.0862 seconds; t=2.229, p=0.0266] **(Figure 2C**), and gaze variability was greater for ASD than the TD group for the robot condition [TD: 0.0054 ± 0.00083 s.d.; ASD: 0.0163 ± 0.0049 s.d.; t=-2.7223, p=0.0236] (**Figure 2D**).

**Figure 2.**
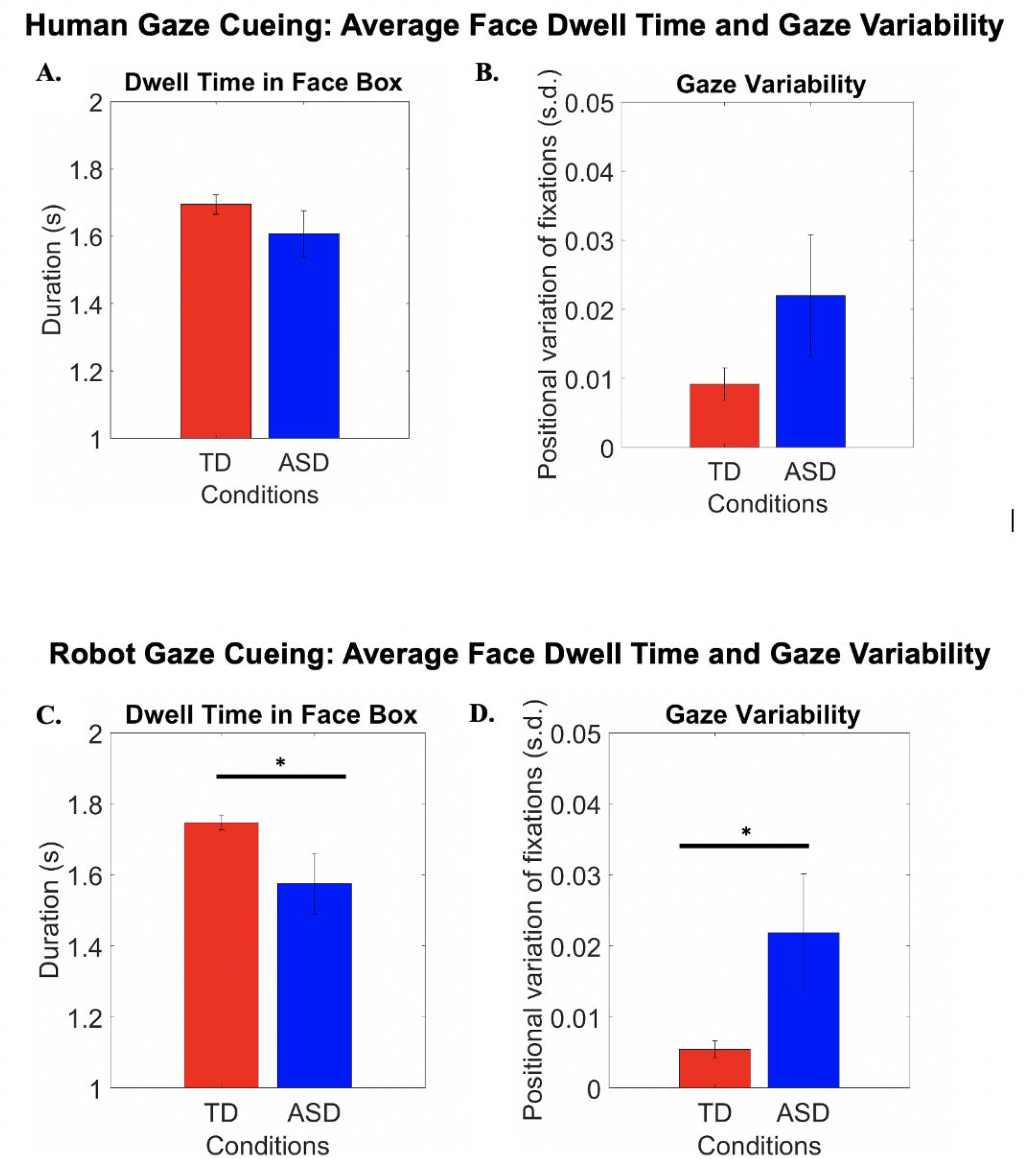
**(A)** Duration of time spent on the face for TD (red) and ASD (blue) participants during the first 2-seconds of the Human Gaze Cueing conditions. **(B)** Standard deviation of horizontal gaze trajectory normalized by duration of face contact. **(C)** Duration of time spent on the face for TD (red) and ASD (blue) participants during the first 2-seconds of the Robot Gaze Cueing condition. **(D)** Standard deviation of horizontal gaze trajectory normalized by duration of robot face contact. Error bars show SEM. *p≤0.05.

### Modulation of Face-Processing Neural Circuitry by Dwell Time

To determine how visual sensing mechanisms modulate neural processing of robot and human faces, neural responses during each two second eye viewing period were modulated by dwell time for both TD and ASD groups. The dwell time variable used in the first level analysis was constructed by identifying gaze duration spent on the face for each two second period prior to changes in gaze direction. In the TD group, the human gaze cueing condition elicited increased activity in the superior parietal lobule including the supramarginal gyrus (SMG) and somatosensory association cortex (SSAC) (**Figure 3A**). On the other hand, the ASD group did not show increased hemodynamic activity in the right superior parietal lobule, but instead showed increased frontal activity (**Figure 3B**). During robot gaze cueing, TD participants showed increased activity in rTPJ and frontal areas (**Supplementary Figure 2**). ASD participants showed increased activity in the superior parietal lobule, motor, and frontal areas (**Supplementary Figure 2**).

**Figure 3:**
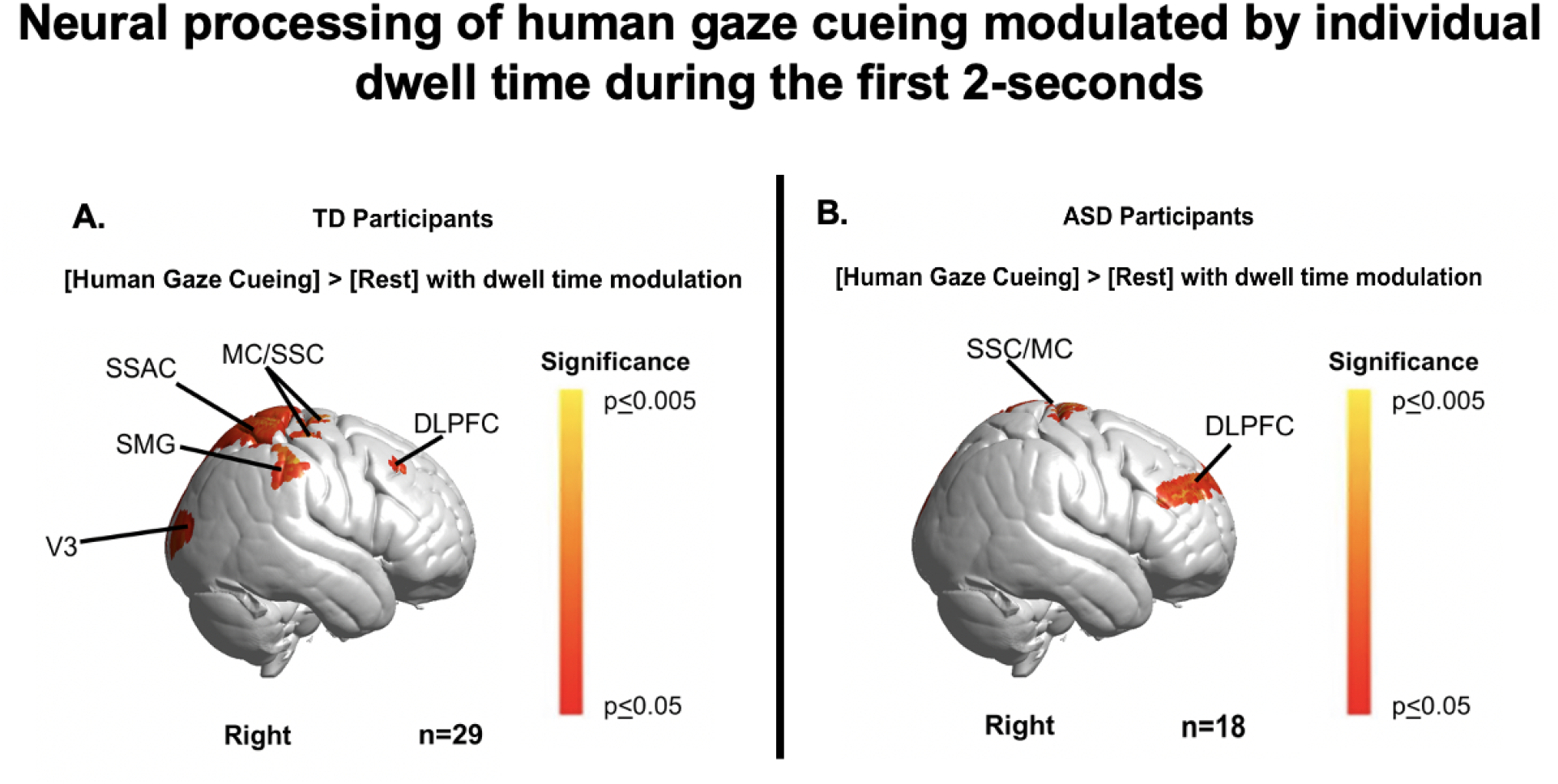
Contrast comparison [Human Gaze Cueing] > Rest] modulated by the duration spent on the human partner’s face within each 2-second eye viewing period. **(A)** TD participants recruited supramarginal gyrus (SMG); somatosensory association cortex (SSAC); dorsolateral prefrontal cortex (DLPFC); frontal eye fields (FEF); pre- and supplementary motor cortex (MC); and extrastriate visual cortex (V3). See **Table 2. (B)** ASD participants showed activation in SSC, MC, and DLPFC. Note: Eye tracking could not be acquired on one TD participant and two ASD participants. See **Table 3**.

### Neural responses during Gaze Cueing modulated by Autism Symptom Severity

Beta values (model-based fNIRS signals) acquired during human gaze cueing were modulated by individual BOSA scores using the General Linear Model (GLM). Blue clusters indicate cortical areas where neural activity is related to individual BOSA scores lower than the ASD participant average. Average BOSA scores were 7 ± 0.42. Here, ASD participants with higher BOSA scores and greater symptomatology showed decreased gaze cueing-related neural activity in the SMG, SSC, FEF, and frontal polar cortex (FPC) (**Figure 4**). (Seven of the 20 ASD participants only had ADOS and not BOSA scores.) The individual BOSA scores for each participant (x-axis, **Figure 5**) are plotted against fNIRS median beta values (y-axis, **Figure 5**) for the SMG and SSC region of interest. The negative correlation, r = −0.86, supports a symptom-related link between the right superior parietal lobule (SMG and SSAC) activity and live human face processing.

**Table 2.**
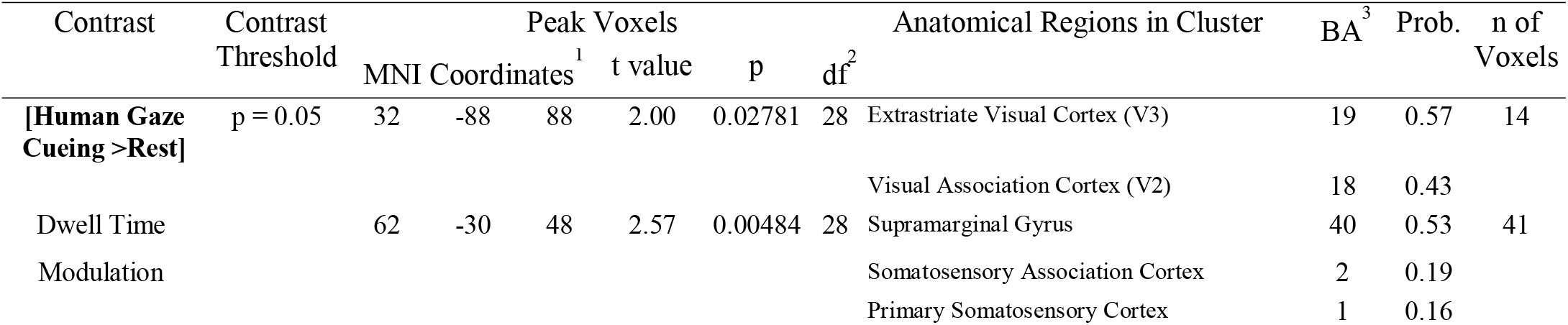
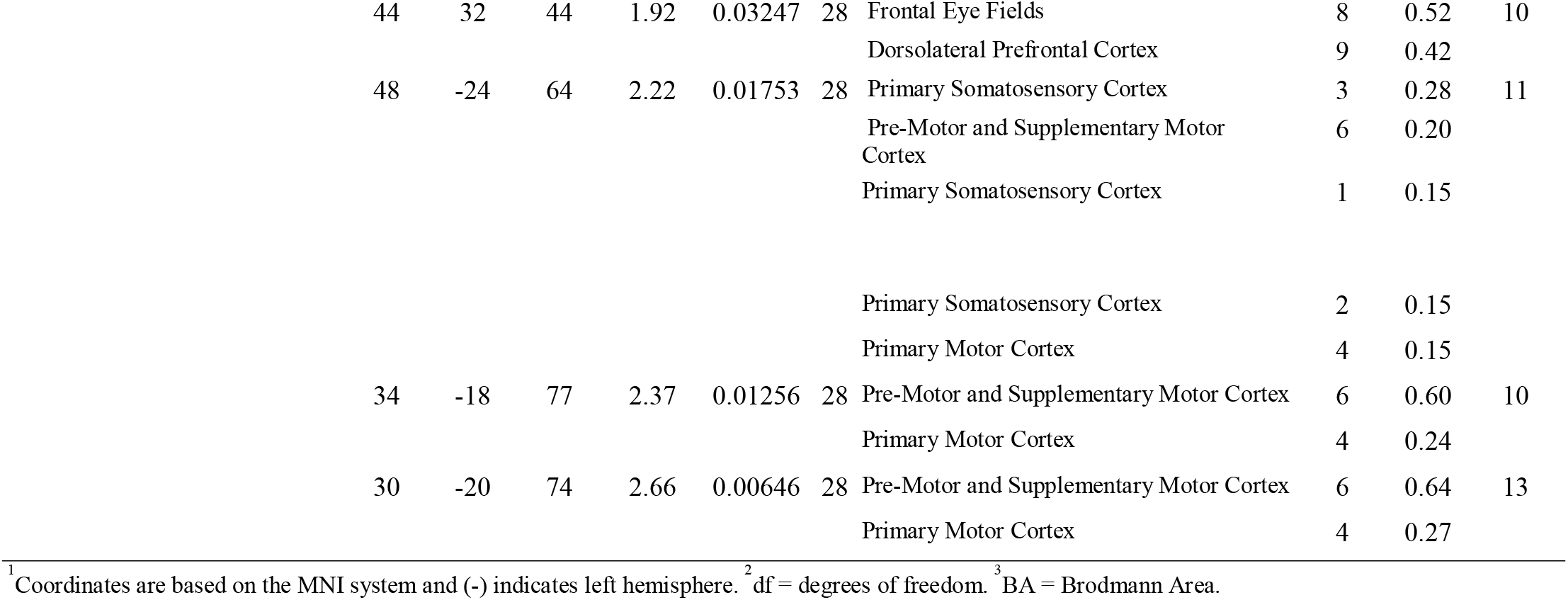
GLM Contrast comparison: [Human Gaze Cueing] > [Rest] modulated by dwell time (deOxyHb + OxyHb signals), TD group

**Table 3.**
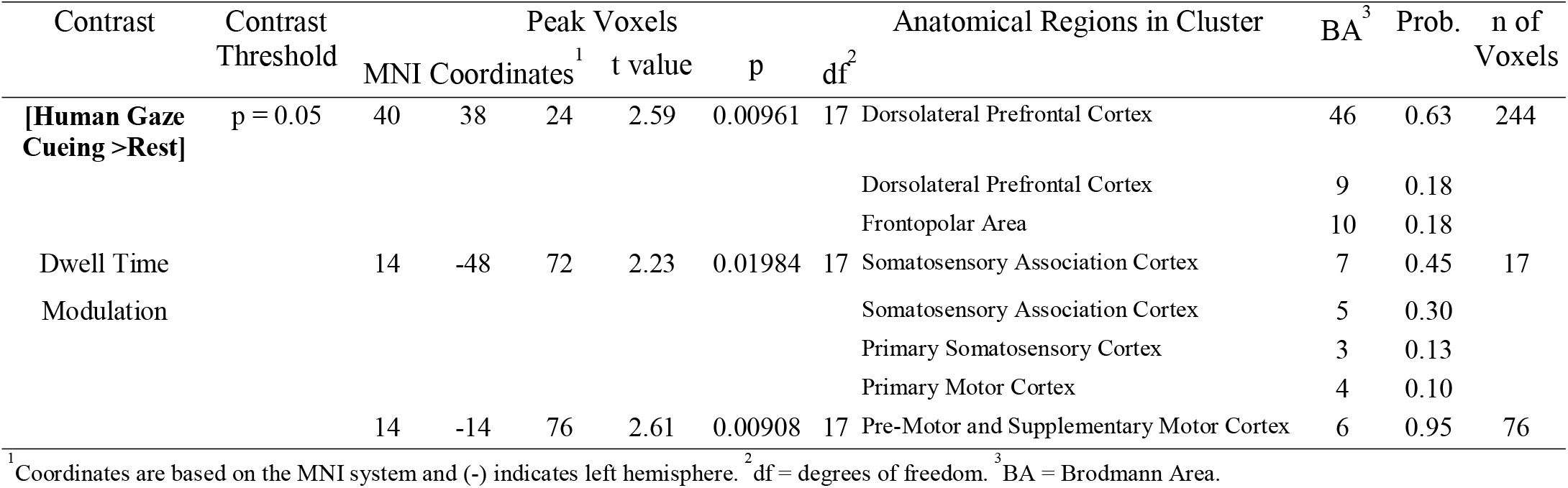
GLM Contrast comparison: [Human Gaze Cueing] > [Rest] modulated by dwell time (deOxyHb + OxyHb signals), ASD group

**Figure 4.**
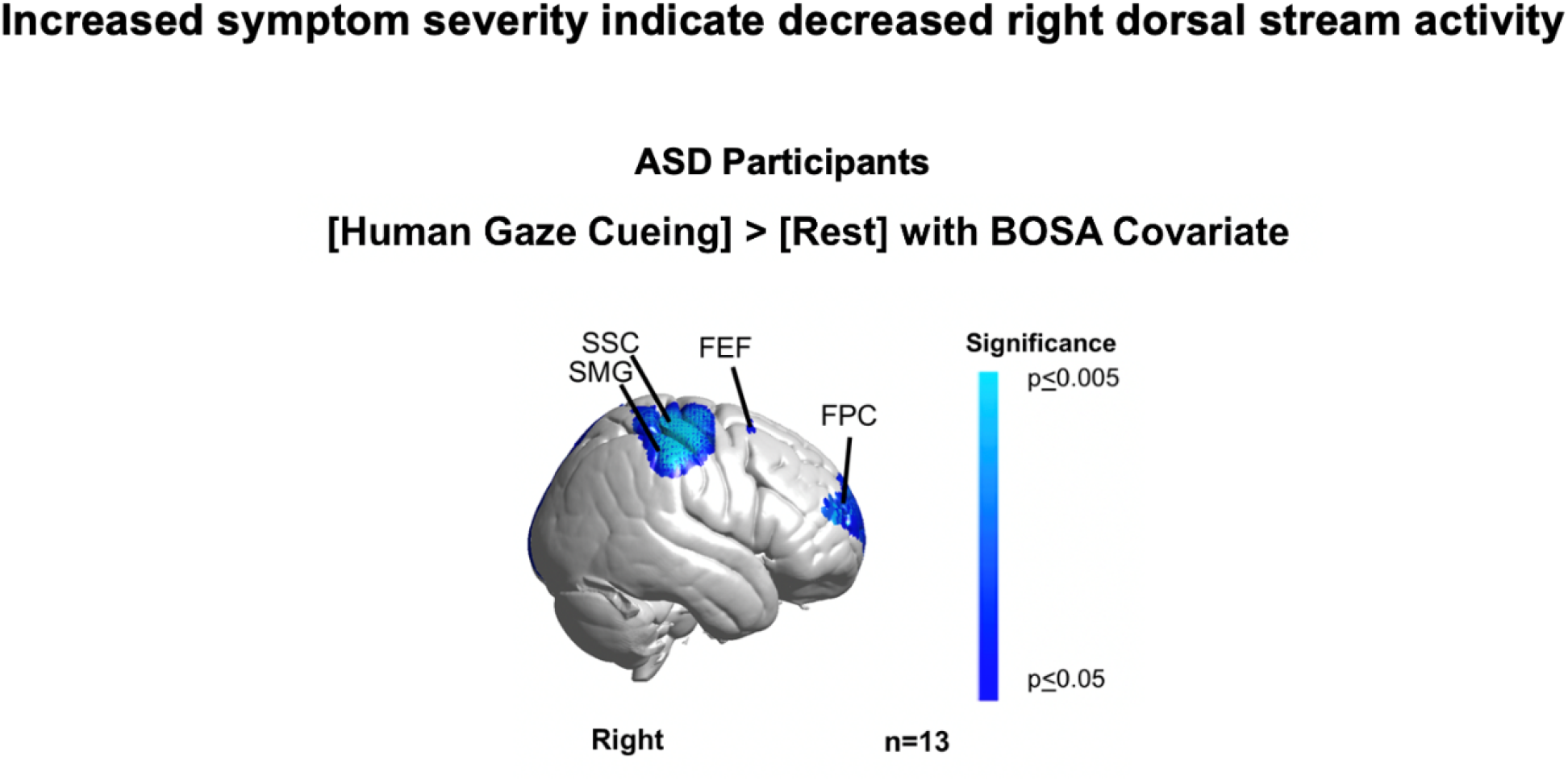
Main effect neural results for Autism Spectrum Disorder (ASD) and typically developed (TD) participants with BOSA-2 (Brief Observations of Symptoms of Autism) scores as a second-level covariate. Blue colors demonstrate a negative relationship between neural responses and BOSA-2 scores, which suggests that increased symptom severity is associated with reduced regional neural responsiveness. Light blue represents responses at p<0.005. FEF: frontal eye fields; SMG: supramarginal gyrus; SSC: somatosensory cortex; and FPC: frontal polar cortex. Seven out of the 20 ASD participants were not assessed using the BOSA scores since they had ADOS scores less than three years old. See **Table 4**.

**Figure 5.**
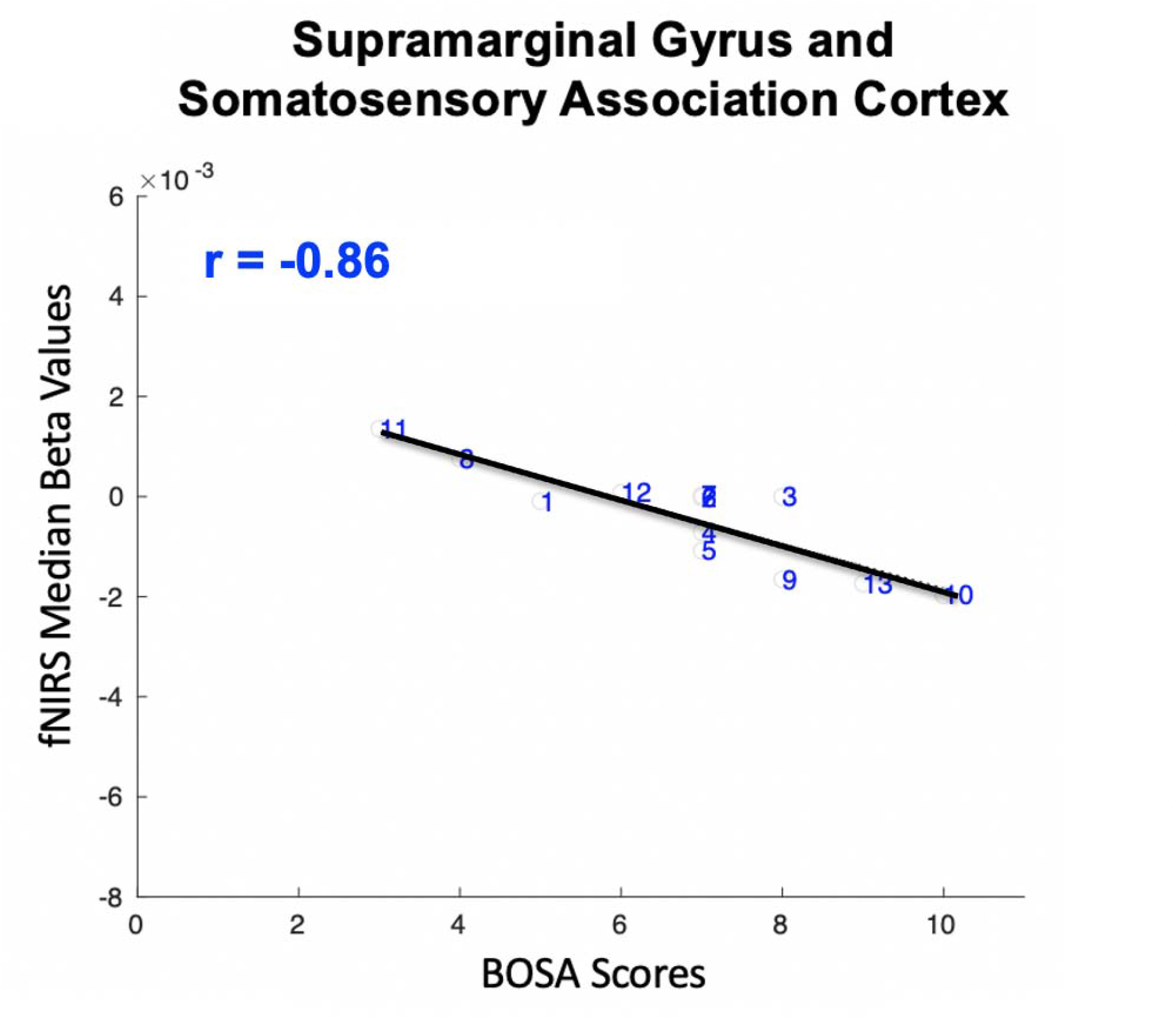
Autism Spectrum Disorder (ASD) participants during human gaze cueing: Median Beta Values vs. BOSA (Brief Observation of Symptoms of Autism) scores. The median beta values (y-axis) within the supramarginal gyrus and somatosensory association cortex (**Figure 4**) and BOSA scores (x-axis) are shown for each ASD participant. The main effect of human gaze cueing is negatively correlated with fNIRS signals in right somatosensory association and supramarginal gyrus cortex (r = −0.86).

To further test if neural encoding of facial information to the dorsal parietal circuit is uniquely affected by live human faces, we examined the dorsal parietal stream and autism symptom severity during the processing of robot faces. Red clusters indicate neural activity in brain areas that are positively related to the individual BOSA scores. Increased neural activity in the SMG, SSC, MC, DLPFC, and FPC during robot gaze cueing was seen regardless of BOSA score (**Figure 6**). The individual BOSA scores for each participant (x-axis, **Figure 7**) are plotted against fNIRS median beta values (y-axis, **Figure 7**) for SMG and SSC and provide no evidence of a correlation between neural activity and autism symptom severity. These findings suggest that dorsal parietal activity is not compromised while following the eyes of robot faces in ASD (regardless of symptom severity).

**Table 4.**
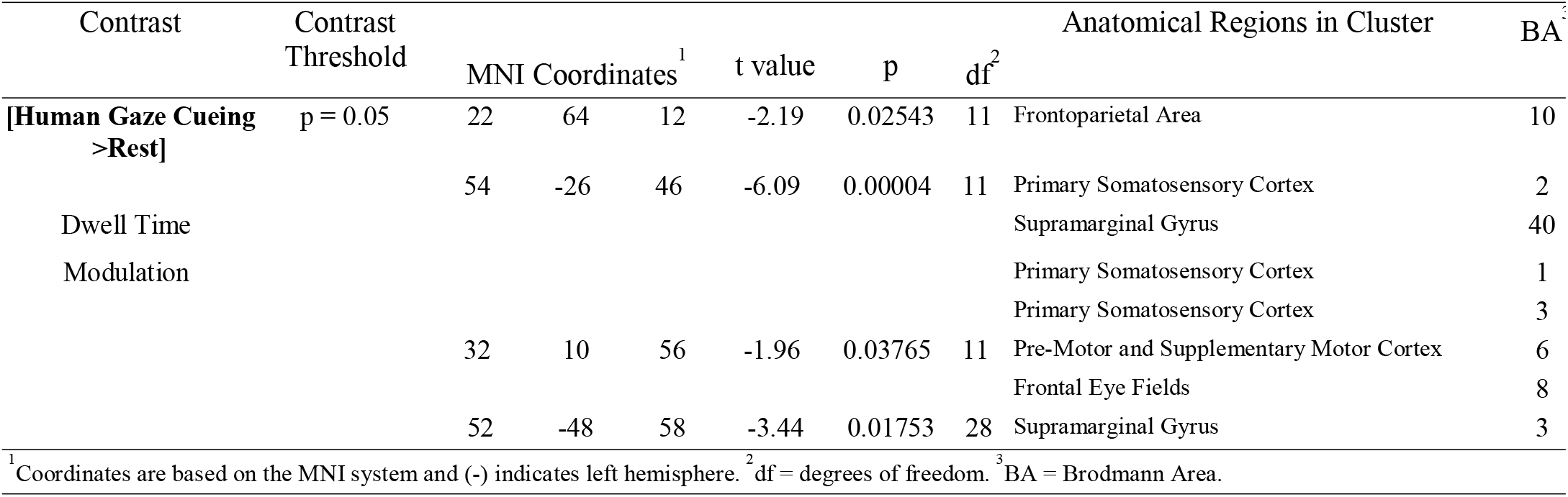
GLM Contrast comparison: [Human Gaze Cueing] > [Rest] with BOSA Covariate (deOxyHb + OxyHb signals), AS

**Figure 6.**
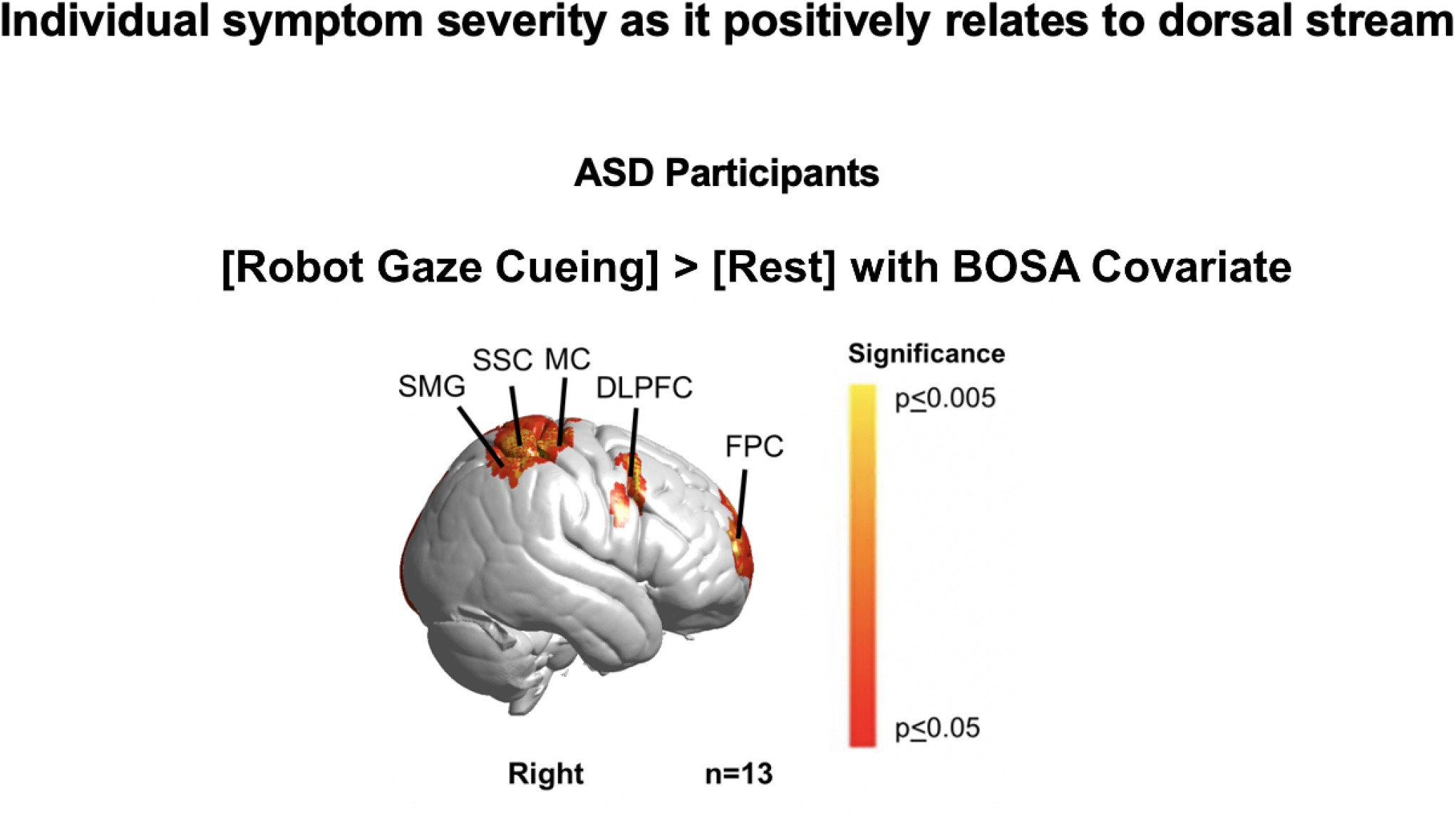
Neural responses for ASD participants with BOSA-2 (Brief Observations of Symptoms of Autism) scores as a second-level covariate. Red colors indicate a positive relationship between neural responses and BOSA-2 scores indicating as symptom severity increases, neural responsiveness in these areas increases. Yellow indicates responses corrected for multiple comparisons at p≤ 0.005. MC: pre-supplementary motor cortex; DLPFC: dorsolateral prefrontal cortex; SMG: supramarginal gyrus; SSC: somatosensory cortex; and FPC: frontal polar cortex. Seven out of the 20 ASD participants were not assessed using the BOSA scores since they had ADOS scores less than three years old. See Table 5.

**Figure 7.**
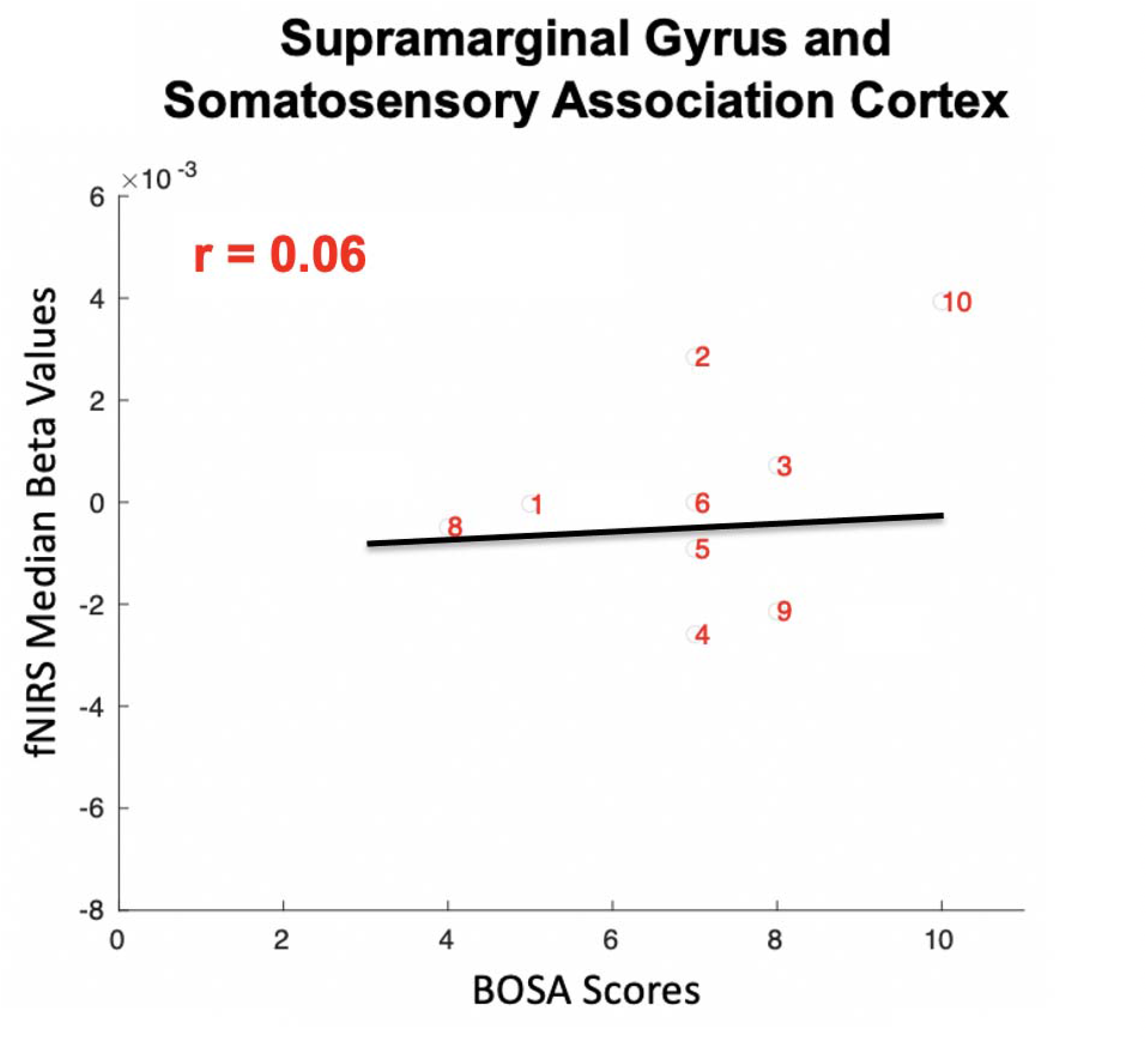
Autism Spectrum Disorder (ASD) participants during robot gaze cueing: Median Beta Values vs. BOSA (Brief Observation of Symptoms of Autism) scores. The median beta values (y-axis) within the supramarginal gyrus and somatosensory association cortex (**Figure 6**) and BOSA scores (x-axis) are shown for each ASD participant. The main effect of human gaze cueing is minimally positively correlated with fNIRS signals in right somatosensory association and supramarginal gyrus cortex (r = 0.06). Due to variations in optode coverage over the supramarginal gyrus and somatosensory association cortex, only ten participants had sufficient data for this regional analysis.

## Discussion

Neural measurements during the interactive social behavior of ASD adults in natural settings holds promise to clarify the neural underpinnings of social behaviors and experiences in ASD. The objective of this study was to examine the neural and visual sensing mechanisms involved in live gaze-directed joint attention in ASD and the relationship to symptomatology. A hallmark of ASD is a difficulty in following eye-gaze during joint attention situations. In this study, we consider that the ability to engage in complex social behaviors, such as eye following, is associated with decoding and initiating social cues for joint and shared attention (Dawson et al., 2004).

**Table 5.**
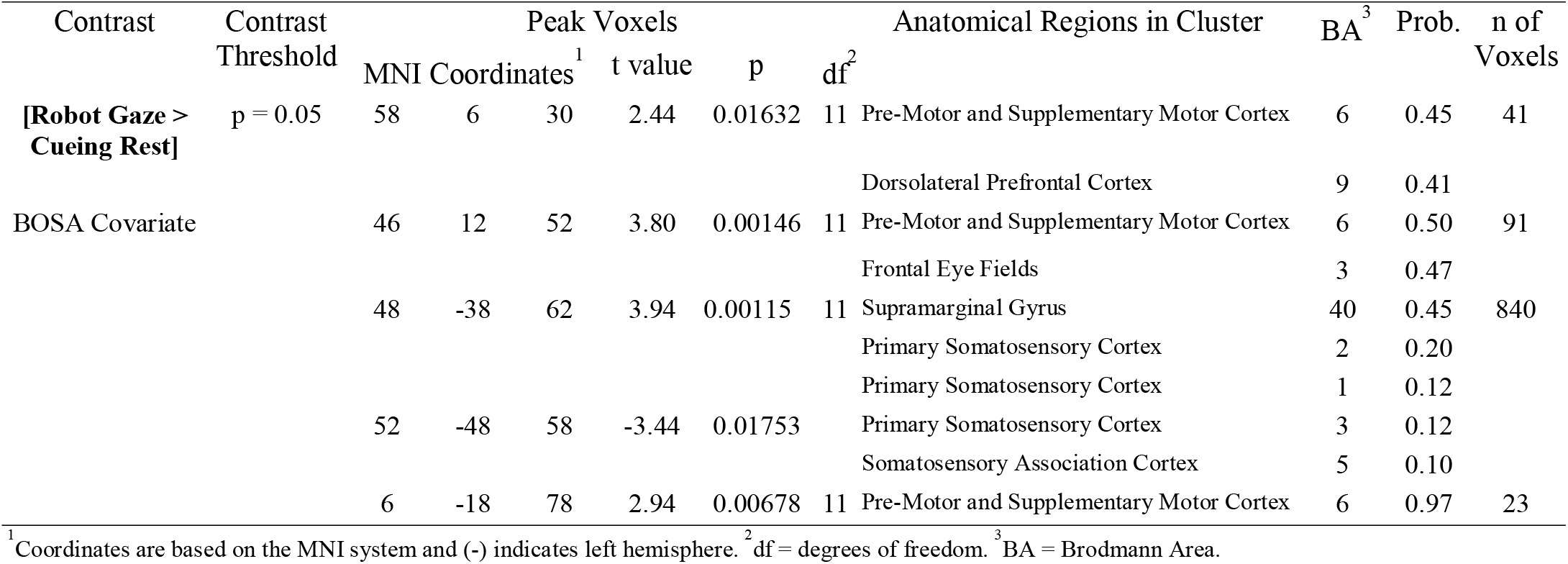
GLM Contrast comparison: [Robot Gaze Cueing] > [Rest] with BOSA Covariate (deOxyHb + OxyHb signals), ASD group

### Visual Sensing Behavior

The use of eye-movement recordings to measure gaze following has the potential to reveal subtle differences in variations in oculomotor and visual sensing mechanisms in ASD. The findings indicate that the ASD and TD groups performed similarly for human-directed gaze and differently for robot-directed gaze. There were no significant differences in the duration spent on the human face, suggesting that ASD individuals gathered sufficient information from the face to shift their attention to the targeted stimulus. There were no significant differences in positional variability to the human face, although the trend was similar to previous results (Hirsch et al., 2022). On the other hand, ASD individuals spent a shorter time looking at the robot face and showed increased positional variability, indicating that ASD individuals use visual sensing mechanisms differently than TD individuals when gathering information from a simulated social face. This particular finding may contribute to the body of evidence that ASD individuals process cartoon-like faces better than human faces (Cross et al., 2022; Rosset et al., 2010).

### Neural Responses modulated by Visual Sensing

We compared how live face information was processed in autistic and non-autistic brains. Consistent with the model of an alternative live face gaze system (Hirsch et al., 2022), human gaze cueing resulted in increased activity of ventral frontal systems in the ASD brain. Specifically, neural findings modulated by human face dwell time revealed increased right dorsal parietal activity for TD participants and increased dorsolateral prefrontal activity for ASD participants. Taken together, these results are consistent with the hypothesis of hypoactive dorsal parietal live eye processing systems in ASD. However, neural responses modulated by robot dwell time revealed that the dorsal parietal lobule and frontal systems are activated by robot gaze in ASD participants. These results suggest that ASD participants acquire features from the robot face to recruit the typical dorsal parietal system. Considering that the dorsal parietal system is seen as a specialized neural circuit for live human eye gaze (Dravida et al., 2020; Hirsch et al., 2022; Kelley et al., 2021; Noah et al., 2020), these findings may add to the existing evidence that robot gaze may be more socially salient in the autistic brain (compared to the typical brain). Similar to the conclusions drawn from previous research on cartoon faces, hierarchical specialization of different types of faces may differ between TD and ASD individuals.

### Neural Signal Association to System Severity

Two large-scale patterns of neural activity were correlated with symptom severity in the BOSA scale. During live human gaze cueing, increased BOSA scores (higher levels of ASD symptomatology) resulted in decreased right dorsal parietal activity, suggesting that encoding of incoming human face information in the dorsal parietal stream may be related to autism symptom severity. During robot gaze cueing, however, dorsal parietal and frontal activity did not relate to autism symptom severity. These correlational findings connecting the neural signal to behavior support the model of alternative live processing of social and simulated social gaze cueing in ASD. Furthermore, the association between neural activity and clinician-reported social function suggests the possibility of a biomarker for ASD (McPartland, 2017).

### Hierarchical Model of Real-Life Interactive Face Processing

The results are consistent with the hierarchical model of real-life interactive face processing. Visual sensing involves the active sampling of a given scene using a systematic pattern of eye movements and fixations (Schroeder et al., 2010). The perceptual cues that signal attention and the visual acquisition of these cues promote joint attention (Mundy & Sigman, 1989). These spontaneous and flexible skills in TD individuals happen early in life, with typical children directing others’ attention by one year (Meltzoff & Moore, 1999). This study includes adults ages 18-40, and thus the major developmental periods for these skills have already passed. However, our results indicate that atypical neural processing of gaze cueing in ASD affect joint attention and social cognitive skills in adulthood.

Our findings are among an increasing number of findings that report instances of alternative neural processing systems for attention to faces and eyes in the context of dynamic face-to-face interactions (Hirsch et al., 2022 2021; Jung et al., 2016) and hypoactivity of the rTPJ system (Redcay et al., 2013). Although various studies have examined whether the patterns of face gaze differ in people with ASD (Pantelis & Kennedy, 2017; Rigby et al., 2016), the neural mechanisms in live gaze cueing in ASD have been understudied. The aim to understand the neural mechanisms that underlie social function in ASD has motivated the multimodal application of fNIRS and eye tracking to compare responses to live human initiators and robot initiators.

## Limitations

There are several limitations to consider for the present study. First, although fNIRS allows for the exploration of live two-person social interactions, it is unable to capture subcortical neural responses that may differ between the two diagnostic groups. Further, only 4% of the participants (6.67% in the TD group and 0% in the ASD group) in the current study were Black when the Black population of New Haven, CT area is 33% (Census, 2020). African, African American, Caribbean, and people mixed with these ethnicities are often excluded from fNIRS studies because of phenotypic factors such as hair-type (Loussouarn et al., 2007; Ricard et al., 2022; Takahashi, 2019). Therefore, future studies must incorporate community-based methods for autism research (Maye et al., 2021) and develop inclusive fNIRS methods (Girolamo et al., 2022; Parker & Ricard, 2022) to recruit samples with a more diverse racial and ethnic representation.

## Conclusion

In conclusion, these findings highlight that altered encoding of live human face information to the superior parietal lobule is a topic of interest for understanding the neural mechanisms governing live joint attention in ASD. The specificity of these findings opens new directions for investigating the connection between the neural encoding of live faces and ASD symptomatology. Our results are consistent with three possible causes for differences in the neural signal in the autistic brain: 1) gating of incoming face information to the dorsal processing stream, 2) lack of acquisition of low-level face features necessary to recruit the dorsal processing stream, and 3) inability to process interactive-specific signals that convey reciprocal social cues. Future research may use techniques such as granger causality (Ono et al., 2022), functional connectivity based on psychophysiological interactions (PPI) analyses, and neural coupling (Hirsch et al., 2022) to study alternative circuitry in the autistic brain during live interactive human face processing. Neural findings of robot gaze cueing are consistent with the hypothesis that hypoactive encoding of incoming information to the dorsal parietal cortex in ASD is specific to live human faces. Based on these results, future research to investigate the therapeutic use of real-life robots in improving dorsal parietal plasticity is suggested.

## Data Availability

All data reported are available on a publicly accessible data repository, DRYAD (www.datadryad.org) at https://doi.org/10.5061/dryad.p5hqbzkt4.

## Acknowledgments

The content of this report is solely the responsibility of the authors and does not necessarily represent the official views of the National Institutes of Health. The authors are grateful to the participants for their essential efforts to advance the understanding of ASD. The authors are also grateful to our two lab partners, JB and TB, for consistent partnership with our participants and the investigators. The authors would also like to thank Jen Cuzzocreo, Erin MacDonnell, and Bela Ponjevic for participant recruitment and database management. Lastly, we would like to thank Christine Curkar-Capizzi and Julie Wolf from the McPartland Lab and Yale Developmental Disabilities Clinic for performing clinical evaluations on the ASD participants. This work was supported by NIH awards NIMH R01 MH111629 (JH and JM), NIMH R21 MH113955 (JH), and NIMH107426 (JH), NSF GRFP #1752143 (TCP).

## Disclosure of Biomedical Financial Interests and Potential Conflicts of Interest

The authors declare that this research was conducted in the absence of any commercial or financial relationships that could be construed as a potential conflict of interest.

## Data Availability Statement

All data reported are available on a publicly accessible data repository, DRYAD, Dataset, https://doi.org/10.5061/dryad.p5hqbzkt4.

## Supporting Information

### Supplementary Methods

#### Participants

Both ASD and TD participants were characterized by self-reported responses from the Social Responsiveness Scale (SRS-2 (Constantino & Todd, 2003)) were reported. The SRS-2 is a quantitative measure of an individual’s impairment related to ASD over the past six months. The SRS-2 consists of five subscales that measure social awareness, social cognition, social motivation, social communication, restricted interests, and repetitive behaviors. Each statement is rated on a four-point Likert scale from 1 (not true) to 4 (almost always true). Higher total scores correspond to greater adversity in socialization. Raw scores of this questionnaire did not differ statistically between diagnostic groups [TD: 56 ± 2.43; ASD: 67 ± 8.3180; t=-1.4804; p=0.1453].

#### Functional NIRS Signal Acquisition, Channel Localization

Hemodynamic signals are acquired using the whole head fNIRS Shimadzu LABNIRS system located in the Brain Function Laboratory. Each emitter transmitted three wavelengths of light, 780, 805, and 830 nm, and each detector measured the amount of light that was not absorbed. The amount of light absorbed was then converted to concentrations of OxyHb and deOxyHb using the modified Beer-Lambert equation. The temporal resolution of fNIRS signals is 27 ms sampling periods with a spatial resolution of 3 cm. Acquired NIRS signals are synchronized with simultaneous eye-tracking. Synchronizing triggers use a TTL pulse relating to the time series of eye-tracking and NIRS signals. Functional NIRS optodes consisted of 40 emitters and 40 detectors arranged in a custom matrix, providing a total of 134 acquisition channels per participant. For consistency, the placement of the most anterior channel of the optode holder cap was centered 1 cm above nasion. To assure acceptable signal-to-noise ratios, resistance was measured for each channel before recording, and adjustments were made for each channel until all recording optodes were calibrated and able to sense known quantities of light from each laser wavelength (Noah et al., 2015; Ono et al., 2014; Tachibana et al., 2011). After the experiment, a 3D Scanner (Structure.IO) was used to record the position of the fNIRS optodes, as well as five anatomical locations (nasion, inion, Cz, left tragus, and right tragus) for each participant (Eggebrecht et al., 2014; Eggebrecht et al., 2012; Ferradal et al., 2014; Okamoto & Dan, 2005; Singh et al., 2005). Montreal Neurological Institute (MNI) coordinates (Mazziotta et al., 2001) for each channel were obtained using NIRS-SPM software (Ye et al., 2009). Anatomical correlates were estimated with the TD-ICBM152 atlas using WFU PickAtlas (Maldjian et al., 2004).

#### fNIRS Signal Processing

Wavelet detrending provided in NIRS-SPM (Ye et al., 2009) removed fNIRS baseline drift. Global components attributable to blood pressure and other systemic effects (Tachtsidis & Scholkmann, 2016) were removed using a principal component analysis (PCA) spatial global mean filter (Zhang et al., 2017; Zhang et al., 2016) before the general linear model (GLM) analysis. The combined OxyHb and deoxy-Hb signals are reported in the analyses. The second level (group) analysis includes the combined signal averages, in which comparisons were done between conditions. Event epochs were convolved with the hemodynamic response function provided from SPM8 (Penny et al., 2011) and were fit to the signals, providing individual beta values for each participant across conditions.

**Table S1.**
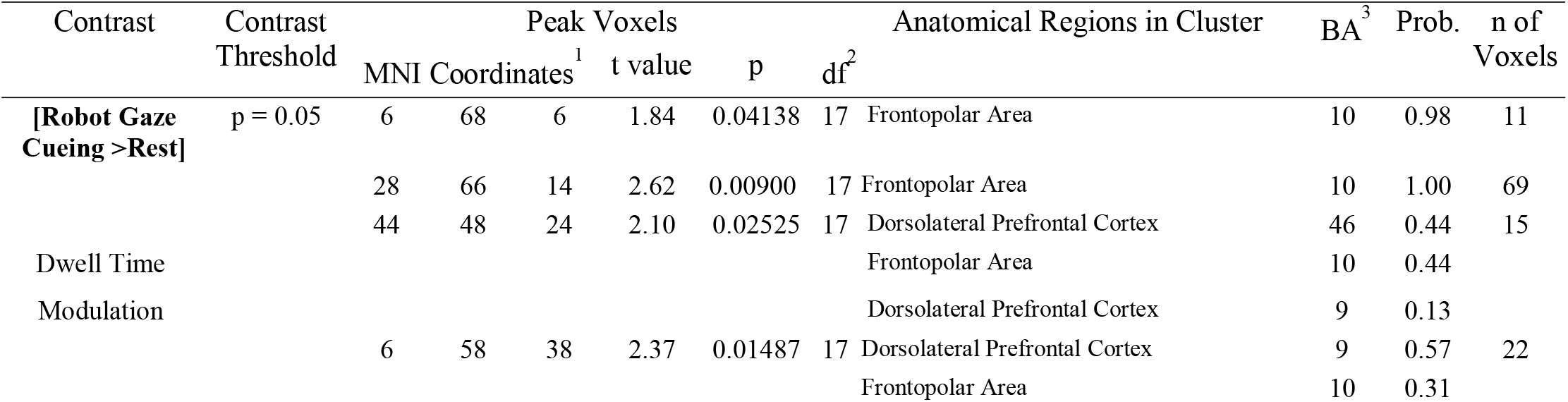
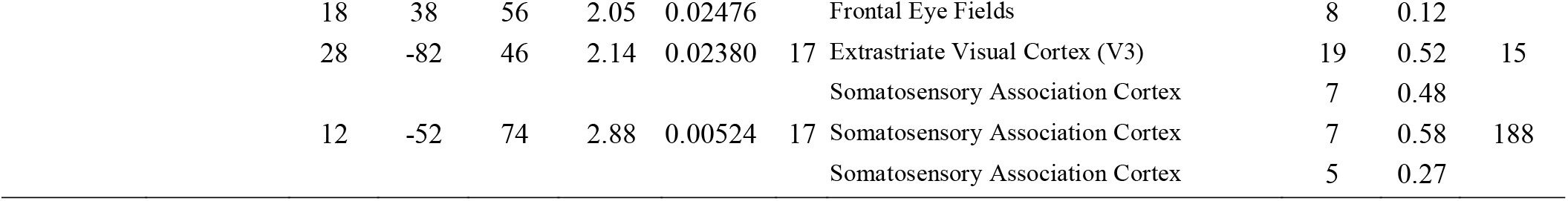
GLM Contrast comparison: [Robot Gaze Cueing] > [Rest] modulated by dwell time (deOxyHb + OxyHb signals), ASD group

**Table S2.**
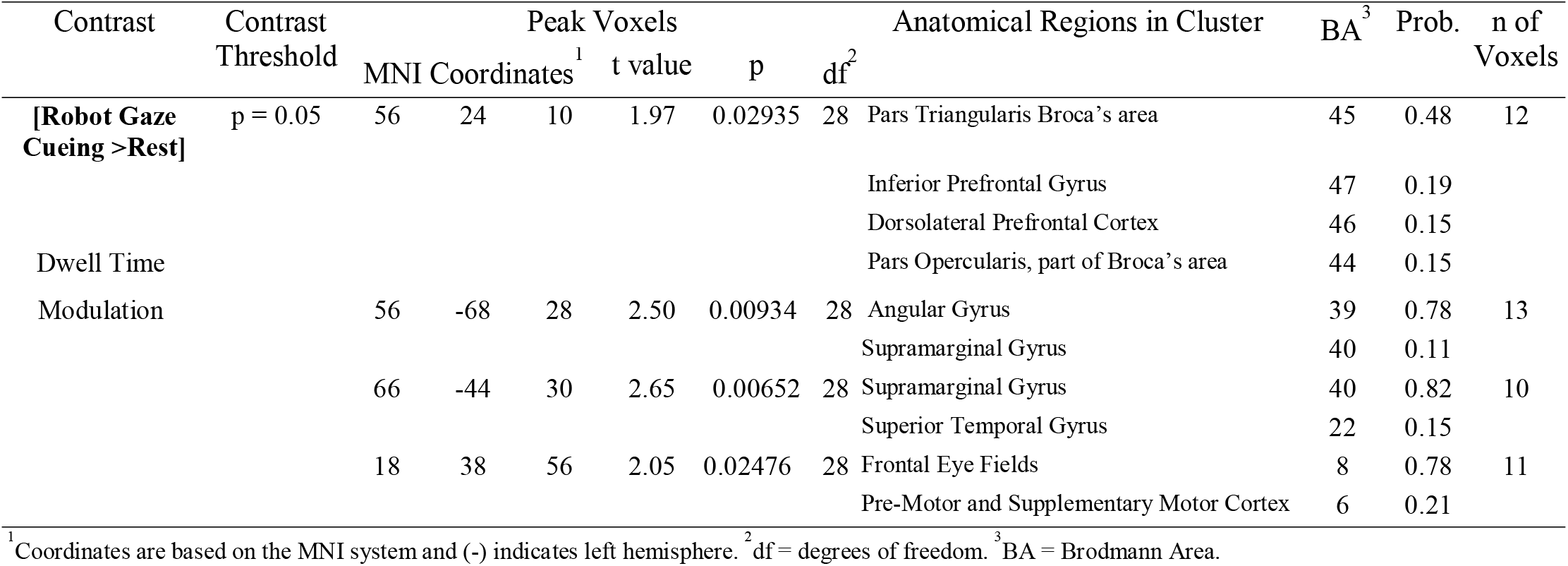
GLM Contrast comparison: [Robot Gaze Cueing] > [Rest] modulated by dwell time (deOxyHb + OxyHb signals), TD group

## Figure Captions

**Supplementary Figure 1.**
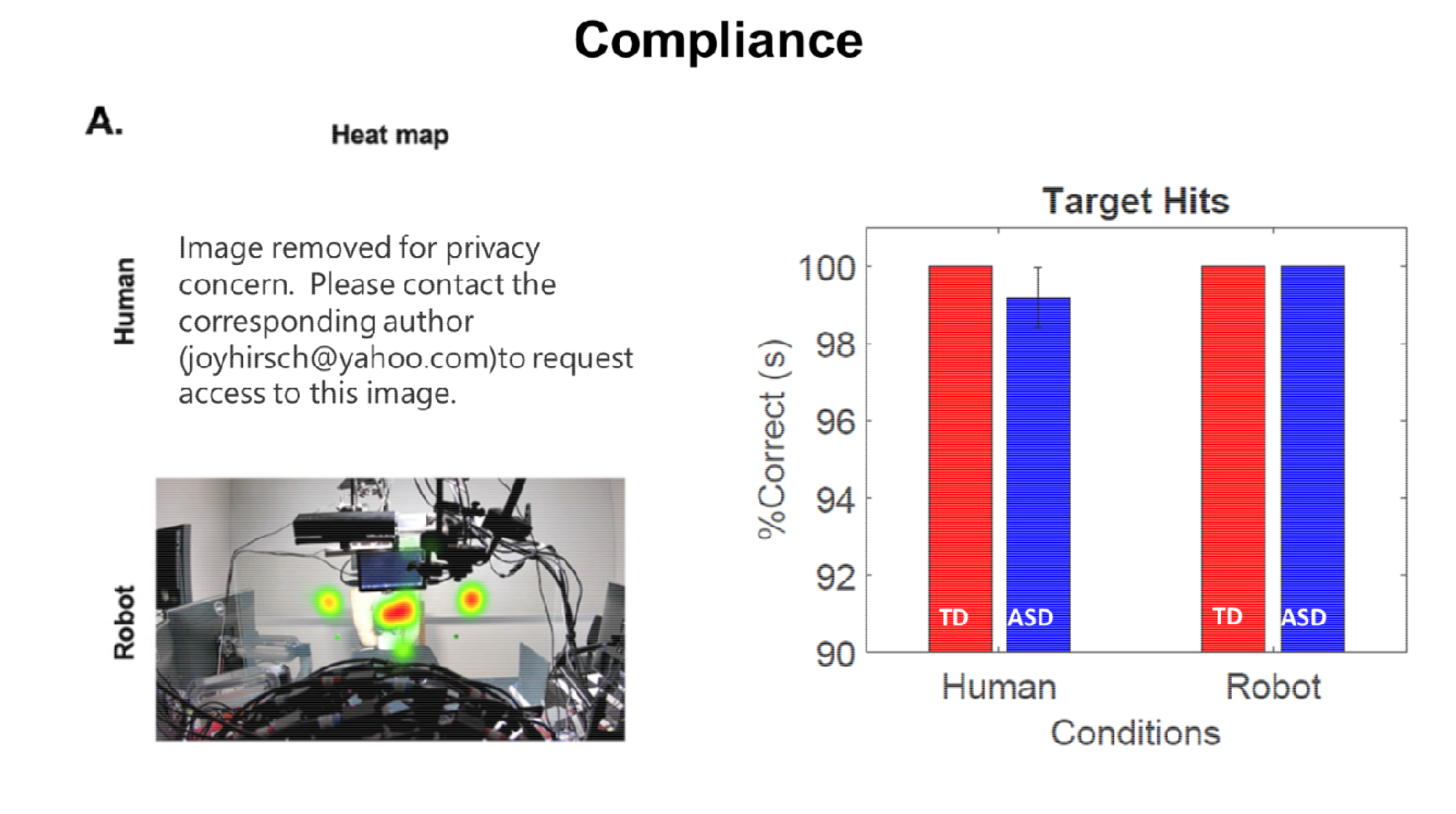
(**A**) Heat maps for Human (top) & Robot (bottom). Eye following trajectories were consistent across conditions. (**B**) Eye-tracking was used to confirm participant compliance for each trial. Blue represents the eye following trajectory of ASD participants and red represents the eye following trajectory of TD participants.

**Supplementary Figure 2.**
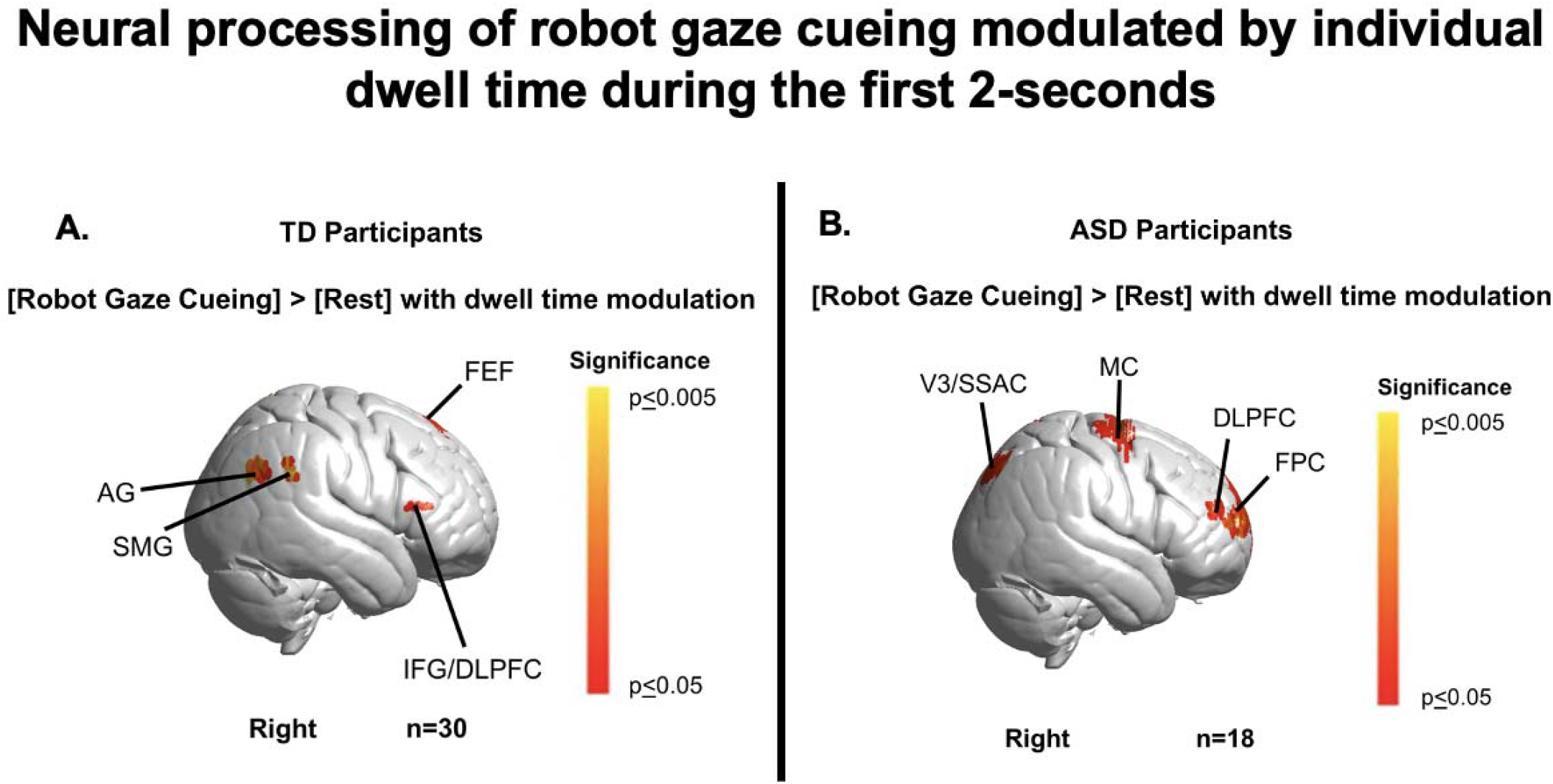
Contrast comparison [Robot Gaze Cueing] > Rest] modulated by the duration spent on the human partner’s face within each 2-second eye viewing period. **(A)** TD participants recruited supramarginal gyrus (SMG); angular gyrus (AG); dorsolateral prefrontal cortex (DLPFC); frontal eye fields (FEF); dorsolateral prefrontal cortex (DLPFC); and inferior frontal gyrus (IFG). See Table S1. **(B)** ASD participants showed activation in frontopolar cortex (FPC); DLPFC; somatosensory association cortex (SSAC); and pre-motor and supplementary motor cortex (MC). Note: Eye tracking could not be acquired on one TD participant and two ASD participants. See Table S2.

